# Microstructural phenotyping and clinical relevance of multiple sclerosis lesions using quantitative susceptibility source separation

**DOI:** 10.64898/2026.10.15.26365462

**Authors:** Eloy Martinez-Heras, Manel Alberich, Hyeong-Geol Shin, Frederic Sampedro, Elvira G-Guzman, Elvira del Agua, Neus Mongay-Ochoa, Jaume Sastre-Garriga, Àlex Rovira, Deborah Pareto

## Abstract

**Background:** Quantitative susceptibility mapping (QSM) exhibits phase cancellation between paramagnetic (e.g. iron) and diamagnetic (e.g. myelin/protein) sources, obscuring microstructural lesion heterogeneity in multiple sclerosis (MS). We evaluated whether biophysical *χ*-separation identifies clinically meaningful lesion phenotypes and whether reduced echo sampling preserves microstructural sensitivity.

**Methods:** In this cross-sectional study of 183 people with multiple sclerosis (PwMS), we analyzed 3327 quality-controlled white matter lesions using 3T MRI with a five-echo gradient-recalled echo sequence (GRE_5_) and a subsampled two-echo dataset (GRE_2_). A hybrid framework combining unsupervised clustering on whole-lesion paramagnetic (*χ*_para_) and diamagnetic (*χ*_dia_) features with rule-based compartmental edge-core gradient assignment resolved MS lesion phenotypes. Intralesional microstructural properties were evaluated using linear mixed-effects models. Multivariable linear regressions assessed associations with physical disability (Expanded Disability Status Scale, EDSS; n = 158), cognitive processing speed (Symbol Digit Modalities Test, SDMT; n = 79), and disease duration (DD; n = 161), with sensitivity models adjusting for total lesion volume, count, or DD.

**Results:** The framework identified four reproducible lesion phenotypes (Adjusted Rand Index = 0.924): Type 1 (22.7%; preserved *χ*_dia_), Type 2 (32.9%; high *χ*_para_ without positive edge-core gradient), Type 3 (35.8%; attenuated *χ*_dia_), and Type 4 (8.6%; high *χ*_para_ with a positive edge-core paramagnetic gradient). Higher EDSS was selectively associated with more negative signed *χ*_dia_ within Type 4 lesions (whole: β = –0.37, *q* = 0.020; core: β = –0.37, *q* = 0.006; edge: β = –0.34, *q* = 0.030) and Type 2 edge (β = –0.33, *q* = 0.044). Type 4 core associations remained robust across sensitivity models adjusting for total lesion volume (β = –0.35, *q* = 0.004), total lesion count (β = –0.37, *q* = 0.008), and DD (β = –0.32, *q* = 0.033). Abbreviated GRE_2_ attenuated Type 4 compartmental associations to non-significance (all *q* greater than 0.05; core: β = –0.20, *q* = 0.215).

**Conclusions:** Quantitative *χ*-separation identified four internally stable susceptibility-defined MS lesion phenotypes, revealing disability-associated tissue heterogeneity beyond total lesion burden. Higher EDSS was associated with more negative diamagnetic measures within Type 4 cores after adjustment for lesion burden, pointing toward complex intralesional non-myelin tissue remodeling or reactive astrogliosis. Two-echo subsampling may influence compartment-specific clinical associations. By uncoupling susceptibility sources, *χ*-separation provides a quantitative framework for characterizing disability-associated lesion heterogeneity.

## 1. Introduction

Multiple Sclerosis (MS) is a chronic, immune-mediated demyelinating disease of the central nervous system characterized by focal inflammatory lesions, diffuse axonal loss, and progressive neurodegeneration^1,2^. Histopathologically, MS white matter (WM) lesions are highly heterogeneous, exhibiting diverse microstructural processes such as different degrees of oligodendrocyte survival, microglial activation and iron deposition^3^. The conceptual framework of MS pathogenesis has undergone a paradigm shift. While acute focal relapses, driven by adaptive immune system infiltrates, dominate the early relapsing-remitting phase of the disease, smoldering neuroinflammation has been identified as the primary driver of non-relapsing, progressive clinical disability^4–6^.

Histopathologically, smoldering neuroinflammation is most characteristically observed in chronic active lesions (CALs). CALs are pathologically defined by a hypocellular, demyelinated core surrounded by a continuously expanding rim of iron-laden, pro-inflammatory microglia and macrophages^7,8^. Recent systematic meta-analyses estimate the prevalence of paramagnetic rim lesions (PRLs) at approximately 52% of PwMS and 12% of lesions, though extreme inter-study heterogeneity exists^9^. Despite this heterogeneity, the presence of PRLs is strongly predictive of earlier conversion to secondary progressive MS and more severe motor disability^10^.

Conventional magnetic resonance imaging (MRI) sequences, such as T2-weighted and T2-fluid-attenuated inversion recovery (FLAIR), are exquisitely sensitive to the macroscopic spatial distribution of MS lesions but suffer from marked pathological ambiguity. They uniformly depict these abnormalities as non-specific hyperintensities, lacking the histopathological specificity to differentiate between acute inflammatory processes and deep axonal destruction. This diagnostic ambiguity is a primary driver of the “clinico-radiological paradox”^11^. In an attempt to bridge this gap, new imaging biomarkers such as slowly expanding lesions (SELs) have been proposed to quantify chronic lesion expansion across longitudinal MRI^12,13^. Through longitudinal non-linear deformation fields (Jacobian determinants), SELs have been established as robust *in vivo* imaging correlates of CALs^14^. Recently, it has been demonstrated that SELs lack gadolinium enhancement and exhibit severe microstructural degradation over time^15^. Specifically, SELs are characterized by progressive drops in magnetization transfer ratio (MTR), diffusion anisotropy and multiparametric T1/T2 relaxometry^16–18^, eventually transitioning into persistent T1 “black holes” indicative of irreversible tissue destruction that strongly predicts greater disability^19,20^. Fundamentally, PRLs are neither strictly synonymous with, nor a direct equivalent of a histopathological CAL. Recent integrated imaging analyses reveal only partial co-localization between the two imaging biomarkers^21,22^. While SELs effectively measure geometric volumetric expansion, this geometric displacement alone cannot resolve the underlying biochemical constituents or confirm the presence of an iron-laden microglial rim^23^. Disentangling this chronic inflammatory microenvironment therefore requires quantitative imaging modalities capable of directly characterizing intrinsic tissue magnetic and microstructural properties^24^.

Quantitative Susceptibility Mapping (QSM) addresses this need using the phase information of gradient-recalled echo (GRE) sequences to quantify the bulk magnetic susceptibility (χ) of tissues in vivo^25^. Because different tissue components exhibit distinct magnetic properties, QSM can provide valuable contrast. Specifically, iron stored within ferritin (abundant in activated microglia) is highly paramagnetic (χ > 0), whereas myelin (composed predominantly of lipids and proteins) is strongly diamagnetic (χ < 0)^26^. However, QSM represents the net linear sum of all magnetic sources within a voxel (χ = χ_para_ + χ_dia_). This additive nature creates a fundamental biophysical ambiguity^27^. For example, an MS lesion undergoing concurrent demyelination (loss of diamagnetic myelin, driving a positive shift in bulk χ) and iron clearance (loss of paramagnetic iron, driving a negative shift in bulk χ) can result in mutual susceptibility cancellation^28^. This inherent cancellation effect actively masks profound, destructive pathological alterations. To resolve this ambiguity, susceptibility source separation (χ-separation) was introduced^29^. The clinical utility and quantitative reliability of χ-separation depend entirely on the quality of the underlying MRI data. The recent ISMRM consensus guidelines for clinical QSM strictly recommend the use of multi-echo 3D GRE sequences with at least five echoes^30^. Acquiring five or more echoes provides highly robust exponential signal decay curves required for accurate phase unwrapping, background field removal, dipole inversion, and stable R2* estimation^24^. Despite these explicit recommendations, evaluating the clinical feasibility of abbreviated MRI protocols is imperative for translational research. Large-scale retrospective multi-center trials and legacy longitudinal datasets frequently employ abbreviated GRE protocols, which typically consist of double-echo GRE sequences to save acquisition time^31,32^. While echo reduction is increasingly accepted for basic QSM, its downstream impact on the more mathematically complex χ-separation pipeline remains critically under-investigated.

To address these critical gaps, we aimed to: (1) Discover reproducible MS lesion phenotypes from decomposed susceptibility sources (χ_para_ and χ_dia_) using a hybrid clustering and rule-based assignment framework, and establish their territorial WM distribution; (2) Uncover their clinical relevance by evaluating disease duration (DD) trajectories and independent associations with Expanded Disability Status Scale (EDSS) and Symbol Digit Modalities Test (SDMT); and (3) Determine whether clinically feasible, abbreviated 2-echo protocols (GRE_2_) maintain the susceptibility source fidelity of standard 5-echo acquisitions (GRE_5_) to resolve independent microstructural clinical associations.

## 2. Methods

### 2.1 Participant cohort

This cross-sectional imaging study evaluated people with MS (PwMS) definitively diagnosed according to the 2017 McDonald criteria^2^. From the initial MRI dataset, 21 PwMS were excluded during standardized preprocessing due to severe susceptibility-related MRI issues, such as motion artifacts and phase corruption following high-pass filtering of the raw phase data^28^. After complete quality control, a final cohort of 183 PwMS (with a total of N = 3327 WM lesions) was retained for all analyses. Comprehensive demographic and clinical markers, including age, sex, DD, SDMT and the EDSS scores, were documented at the time of MRI acquisitions (Table 1). The study protocol was approved by the local institutional review board (Vall d’Hebron Hospital Universitari), and all participants provided written informed consent in accordance with the Declaration of Helsinki.

**Table 1.** Demographic, clinical and macrostructural imaging characteristics of the final MS cohort (N=183).

| Characteristics | Summary Statistics | Available data (n, %) |
| --- | --- | --- |
| **Demographic** |  |  |
| Age at MRI (years), Median [IQR] | 50.0 [42.0 - 58.0] | 183 (100%) |
| Sex (Female / Male), n (%) | 140 (76.5%) / 43 (23.5%) | 183 (100%) |
| **Clinical & physical/cognitive disability** |  |  |
| DD from onset (years), Mean ± SD | 17.8 ± 10.5 | 161 (88.0%) |
| Expanded Disability Status Scale (EDSS), Median [IQR] | 2.0 [1.5 - 4.0] | 158 (86.3%) |
| *Mild disability (EDSS ≤ 2.5), n (%)* | 98 (62.0%) | - |
| *Moderate disability (EDSS 3.0 – 5.5), n (%)* | 36 (22.8%) | - |
| *Severe disability (EDSS ≥ 6.0), n (%)* | 24 (15.2%) | - |
| SDMT standardized to 90 seconds, Mean ± SD [Median] | 40.5 ± 13.2 [40.0] | 79 (43.2%) |
| **Macrostructural MRI lesion burden** |  |  |
| Total lesion count per subject, Median [IQR] | 16 [7 – 26] | 183 (100.0%) |
| Total lesion volume per subject (mL), Median [IQR] | 3.81 [1.00 – 12.10] | 183 (100.0%) |

Values are presented as Mean ± Standard Deviation (SD), Median [Interquartile Range, IQR], or count (percentage, %). Percentages for available data are calculated relative to the final analyzed cohort (N = 183). Disease duration was calculated from the date of first clinical symptom/relapse to the date of MRI acquisition. EDSS: Expanded Disability Status Scale; SDMT: Symbol Digit Modalities Test; IQR: interquartile range (25th–75th percentiles).

### 2.2 MRI Acquisition

All neuroimaging was acquired on a SIEMENS Magnetom Prisma 3T scanner (Siemens Healthineers, Erlangen, Germany) with a 64-channel phased-array head/neck coil. The primary susceptibility-sensitive acquisition consisted of a 3D multi-echo GRE sequence. The precise acquisition parameters for the standard GRE_5_ were as follows: Echo Times (TE): 7.62, 12.78, 17.94, 23.10, 28.26 ms, Repetition Time (TR): 35 ms, Flip Angle: 15°, Voxel Size: 1.0 x 1.0 x 1.3 mm^3^ resolution and 120 axial slices providing complete whole-brain coverage (Acquisition time = 6 min 12 s).

To guarantee perfect spatial and temporal pairing for the comparative analysis, the abbreviated GRE_2_ was generated offline by computationally extracting only the first and last echoes of the standard GRE_5_ acquisition (7.62 and 28.26 ms). This methodological approach eliminated any potential confounding factors related to subject motion, physiological noise variations, or subtle scanner drift between separate physical acquisitions, ensuring that any observed quantitative differences were purely a mathematical consequence of the echo reduction in the processing pipeline.

In addition to the multi-echo GRE sequence, standard clinical structural scans were acquired for all participants to facilitate accurate tissue segmentation: a high-resolution 3D T1-weighted Magnetization Prepared Rapid Gradient Echo (MPRAGE) sequence (TR = 2300 ms, TE = 2.98 ms, TI = 900 ms, flip angle = 9°, voxel size = 1.0 mm isotropic) and a 3D Fluid-Attenuated Inversion Recovery (FLAIR) sequence (TR = 5000 ms, TE =395 ms, TI = 1800 ms, voxel size = 1.0 mm isotropic).

### 2.3 Quantitative susceptibility and χ-separation reconstruction

Multi-echo GRE magnitude and phase images were processed in their native acquisition space to obtain QSM, effective transverse relaxation (R *), and susceptibility-source separation maps (χ_para_ and χ_dia_). For both the GRE_5_ and GRE_2_ pipelines, identical mathematical algorithms and parameters were applied. A native-space brain mask was generated from the first-echo magnitude image using SynthStrip^33^. Magnitude and phase images were ordered by echo number and concatenated into four dimensions with parameters extracted directly from BIDS metadata^34^. Phase processing followed a standardized processing pipeline: raw complex phase data were spatially unwrapped using a multi-echo workflow combining ROMEO and 3D Laplacian unwrapping^35,36^. Non-local background magnetic fields were removed using the variable-kernel sophisticated harmonic artifact reduction on phase data (V-SHARP) method to extract the local tissue phase. Bulk magnetic susceptibility (χ) maps were reconstructed from the local field maps via regularized single-step dipole inversion implemented in the SEPIA framework^37^. In parallel, effective R * maps were estimated from the magnitude decay across echo times: for GRE_5_, R2* was computed via a mono-exponential decay fit across all five available echoes^38,39^, whereas for GRE_2_, it was determined using a two-point logarithmic ratio. QSM and R_2_* maps were subsequently integrated into the biophysical χ-separation model to decouple net magnetic susceptibility into voxel-wise paramagnetic (χ_para_) and diamagnetic (χ_dia_) components^29^,^1^. To reduce contamination from venous susceptibility, a deep-learning-based vessel segmentation was applied independently to both susceptibility maps^40^. All output maps were restricted to the native-space brain mask and stored in the native acquisition space.

### 2.4 Conventional MRI preprocessing and automated lesion segmentation

Structural T1-weighted (T1w) and FLAIR MRIs were skull-stripped using SynthStrip^33^ and rigidly co-registered to common anatomical space. MS lesions were automatically segmented from the co-registered T1w and FLAIR MRIs using LST-AI^41^. The resulting binary lesion mask was separated into individual connected components using 26-neighbour connectivity.

To minimize partial-volume effects and measurement noise, lesions smaller than 27 mm^3^ were excluded^42^. Lesion segmentations were used to generate lesion-filled T1w MRI^43^, which were processed with FastSurfer to obtain a whole-brain anatomical parcellation^44^. Brain WM labels were used to generate normal-appearing white matter (NAWM) reference regions. These masks were conservatively eroded to minimize the effects of tissue boundaries and exclude MS lesions. Finally, lesion location was classified by LST-AI into four anatomical territories: periventricular, juxtacortical, subcortical/deep WM, and infratentorial (brainstem and cerebellum).

### 2.5 Registration and core-edge lesion parcellation

Lesion masks, anatomical parcellations, and reference tissue masks were registered to each quantitative susceptibility acquisition (GRE_5_ and GRE_2_). Each lesion was evaluated independently. To characterize spatial heterogeneity, each lesion was segmented into whole-lesion, core, and edge compartments. The edge was defined as a single in-plane voxel layer (1.0 mm) obtained by eroding the lesion mask using a 2D 3×3×1 in-plane structuring kernel, avoiding cross-slice distortion. The core was defined as the surviving central compartment following erosion, and the edge was obtained by subtracting the core from the whole-lesion mask.

Core and edge measurements were considered valid only when both compartments independently met a minimum physical volume threshold of ≥ 10 mm^3^ to prevent partial-volume effects and compartment loss from morphological erosion. For each valid compartment, mean values were extracted from QSM, χ_para_ and χ_dia_, and edge-core gradients were calculated (Δχ_para,edge-core_ = χ_para,edge_ − χ_para,core_), see Figure 1. Identified vessel voxels set to zero were excluded from calculations, and MS lesions failing to reach quantitative thresholds were excluded from compartment-specific analyses (51 lesions failing QC were excluded).

**Figure 1:**
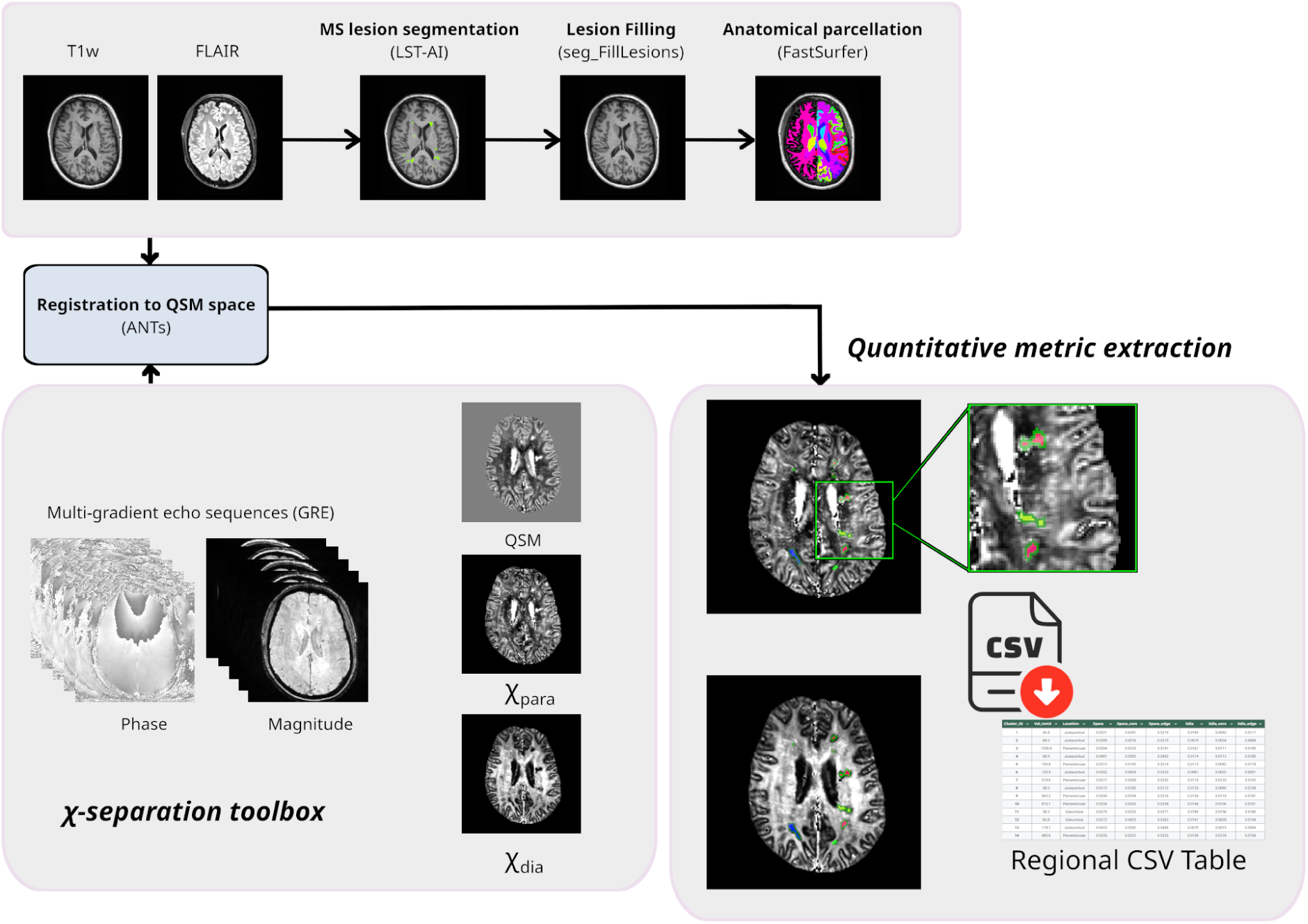
Methodological pipeline for susceptibility-source lesion phenotyping. Overview of the processing framework detailing the co-registration of structural LST-AI lesion masks into native quantitative susceptibility mapping space, followed by 26-neighbor 3D connected component labeling and 1 mm in-plane erosion to parcellate lesions into distinct core and edge compartments. Extracted multimodal metrics (χ, χ_para_, χ_dia_) are fed into an unsupervised hierarchical clustering algorithm to derive microstructural imaging lesion phenotypes.

### 2.6 Statistical and computational framework

All statistical modeling, machine learning analyses, and bootstrap resampling analyses were performed in Python (v3.10) using SciPy, Scikit-learn, and Statsmodels.

#### Hybrid lesion phenotyping and hierarchical assignment

Unsupervised K-means clustering was applied to classify all quality-controlled MS lesions based on whole-lesion susceptibility features (χ_para, whole_, χ_dia, whole_). Primary K partitions (k = 2 to 4) were evaluated using silhouette analysis and Prediction Strength (PS). Intra-cluster feature heterogeneity was subsequently resolved using a two-branch hybrid approach: (1) within the lower-χ_para_ parent cluster, a second data-driven K-means model partitioned lesions according to whole-lesion diamagnetic magnitude (|χ_dia_|); (2) within the higher-χ_para_ parent cluster, a deterministic rule based on a positive edge-core paramagnetic gradient (Δχ_para,edge-core_ > 0 ppb) was applied to segregate lesions with peripheral paramagnetic enrichment from those without a positive gradient. Hierarchical partition stability was assessed using subject-clustered bootstrap resampling distributions (250-500 iterations) of the Adjusted Rand Index (ARI) and per-subtype Jaccard similarity coefficients.

#### Microstructural group comparisons (LMM)

To compare microstructural susceptibility metrics across MS lesion phenotypes while accounting for intra-subject lesion clustering, we employed Linear Mixed Effects Models (LMM) with subject-specific random intercepts. LMMs with subject-specific random intercepts compared whole-lesion χ_para_, displayed diamagnetic magnitude, and diamagnetic deficit Δ|χ_dia_| relative to NAWM across phenotypes, adjusting for log-transformed lesion volume, with multiple-comparison correction using Benjamini-Hochberg False Discovery Rate (BH-FDR *q* < 0.05).

#### Spatial topography modeling

To evaluate territorial predilection across cerebral WM compartments (periventricular, juxtacortical, and subcortical/deep WM) while accounting for intra-subject lesion clustering, multivariable binomial Generalized Estimating Equations (GEE) were used. Lesion phenotype membership was modeled as the outcome, using PwMS as the clustering variable and infratentorial WM as the reference territory.

#### Clinical disability and cognitive association modeling

To evaluate associations with EDSS and SDMT (standardized to 90 seconds), multivariable linear regression models were fitted at the subject level using Ordinary Least Squares (OLS) based on each subject’s mean intralesional susceptibility values per lesion phenotype compartment. Primary models were adjusted for age and sex. To isolate intrinsic microstructural alterations from macroscopic lesion burden, sensitivity models were additionally adjusted for subject-level log-transformed total lesion volume or count. Further models were adjusted for DD to confirm independence from disease stage. Multiple testing was controlled using the BH-FDR *q* < 0.05.

#### Physical disability stratification and lesion phenotype burden escalation

To determine how the MS lesion phenotypes relate to clinical disability, the PwMS cohort with available EDSS scores was stratified into three established disability categories: mild disability (EDSS ≤ 2.5; n = 98), moderate disability (EDSS 3.0–5.5; n = 36), and severe disability (EDSS ≥ 6.0; n = 24). Across these categories, we evaluated: (1) subject-level lesion phenotype prevalence; (2) phenotype-specific macroscopic burden load escalation, comparing median cumulative lesion volumes (mm³, log-scaled); and (3) intralesional susceptibility alterations within the high-χ_para_ parent branch across mild, moderate, and severe disability groups using one-way Analysis of Variance (ANOVA) and multivariable linear regression adjusted for age, sex, and total macroscopic lesion load.

#### Phenotypic composition and DD trajectories

DD from symptom onset was modeled against absolute lesion phenotype burden metrics (log-transformed volumes and counts) using multivariable linear regression adjusted for age and sex. Phenotype volume and count percentages were separately correlated with EDSS using subject-clustered bootstrap Spearman rank correlations (1000 iterations) with 95% bootstrap confidence intervals.

#### Technical protocol validation (GRE_5_ and GRE_2_)

Mean QSM, χ_para_, and χ_dia_ values were extracted from protocol-matched NAWM and the three lesional compartments (whole, core and edge). To evaluate the impact of echo reduction, paired Wilcoxon signed-rank tests, Cohen’s effect sizes for paired differences and Lin concordance correlation coefficients (CCC) were computed comparing GRE_5_ and GRE_2_ across all lesion compartments and across regional anatomical parcellations (Desikan-Killiany cortical and subcortical structures; *q* < 0.05). To evaluate whether abbreviated protocols preserve clinical sensitivity, the entire hybrid phenotyping framework and multivariable clinical models were independently replicated using GRE_2_ data associations.

## 3. Results

### 3.1 Biochemical lesion phenotyping

All primary lesion phenotyping and clinical association analyses were established using the high-fidelity GRE_5_, leveraging multi-echo exponential fitting accuracy for R2* and susceptibility source separation.

Our hybrid clustering and rule-based assignment pipeline partitioned all quality-controlled MS lesions (N=3327 from 183 PwMS) into distinct microstructural susceptibility phenotypes (see Supplementary Figure 1). The primary clustering algorithm (k = 2) achieved high component stability (mean Silhouette Coefficient = 0.40, cross-validated PS = 0.80), segregating the cohort into two broad parent branches relative to the overall lesion cohort median (median χ_para_ = 11.92 ppb): a lower-χ_para_ branch (median 8.83 ppb; n = 1948, 58.6%) and a higher-χ_para_ branch (median 17.73 ppb; n = 1379, 41.4%). To resolve intra-cluster feature heterogeneity, hierarchical assignment was implemented across both branches: (1) within the lower-χ_para_ parent branch, a second data-driven K-means model demonstrated robust cluster separation, partitioning lesions into subclusters based on whole-lesion displayed |χ_dia_| (mean Silhouette Coefficient = 0.41, median Jaccard stability = 0.96); (2) within the higher-χ_para_ parent branch, lesions were deterministically segregated based on an active positive edge-core paramagnetic gradient (Δχ_para,edge-core_>0). Across the full four-phenotype hierarchy, the final MS lesion phenotypes maintained high cross-validated stability (PS = 0.71; bootstrap median ARI = 0.92).

As visualized in Figure 2 and Table 2, these four microstructural profiles are distinctly separated:

● **Type 1 (Preserved diamagnetic susceptibility; N=756, 22.7%):** Derived from the lower-χ_para_ parent cluster, characterized by lowest paramagnetic signal (mean χ_para_= 7.57 ppb) coupled with a strongly preserved diamagnetic signature (χ_dia_= –31.17 ppb; positive magnitude contrast relative to NAWM: Δ|χ_dia_|= +4.12 ppb)
● **Type 2 (Higher paramagnetic susceptibility with a non-positive or unavailable edge-core gradient; N=1094, 32.9%):** Assigned from the higher-χ_para_ parent cluster, characterized by prominent total paramagnetic accumulation (χ_para_= 20.71 ppb), combined with the lowest diamagnetic signal across the cohort (χ_dia_= –17.30 ppb, Δ|χ_dia_| = –9.06 ppb below NAWM), comprising lesions lacking a positive peripheral paramagnetic gradient (including 421 lesions with verified non-positive edge-core gradients and 673 smaller lesions where concentric core-edge erosion did not meet the ≥ 10 mm³ compartment volume threshold).
● **Type 3 (Attenuated diamagnetic susceptibility; N=1192, 35.8%):** Originating from the lower-χ_para_ parent cluster, characterized by low paramagnetic susceptibility below the lesion cohort median (median χ_para_= 9.80 ppb vs cohort median 11.92 ppb), alongside an attenuated diamagnetic signal (χ_dia_= –23.09 ppb; Δ|χ_dia_| = –2.83 ppb below NAWM), denoting a reduction in diamagnetic sources without elevated paramagnetic accumulation.
● **Type 4 (Positive Δχ_para,edge-core_ gradient signature; N=285, 8.6%):** Emerging from the higher-χ_para_ parent cluster (χ_para_= 18.08 ppb), identifying lesions with an invariant positive edge-core paramagnetic gradient (Δχ_para,edge-core_>0; Δχ_para_= +2.89 ppb), marked whole-lesion diamagnetic attenuation (χ_dia_= –19.77 ppb) and a significantly larger median volume (215.8 mm³; Table 2 and Supplementary Figure 2).

**Figure 2:**
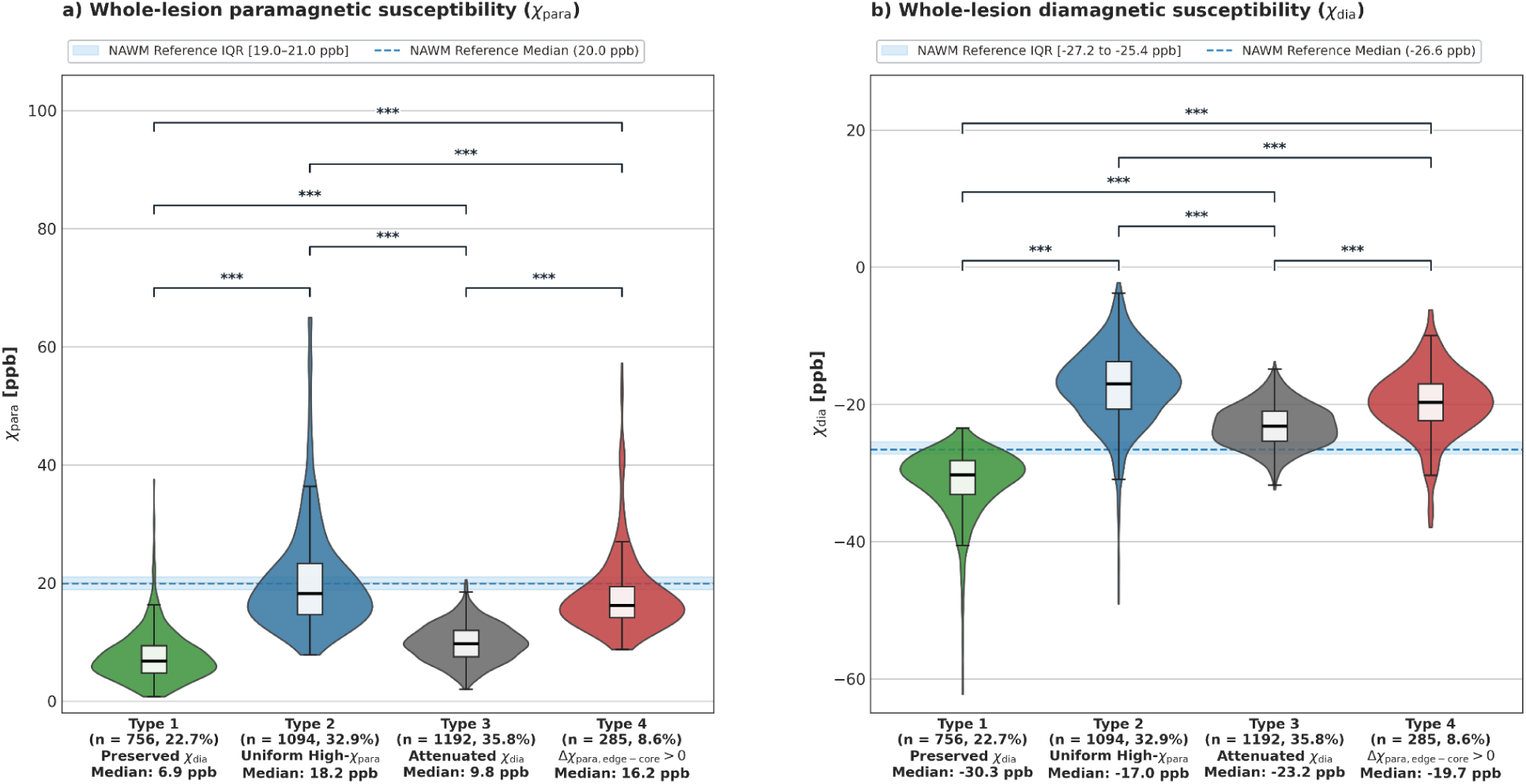
Whole-lesion microstructural susceptibility profiles across MS lesion phenotypes: Distribution of (a) whole-lesion paramagnetic susceptibility (χ_para_) and (b) whole-lesion signed diamagnetic susceptibility (χ_dia_) across 3327 MS lesions from 183 PwMS across Types 1–4. Blue dashed line and shaded band indicate reference median and IQR of normal-appearing white matter (NAWM; χ_para_ = 20.0 ppb, signed χ_dia_ = –26.6 ppb). Brackets denote pairwise LMM contrasts surviving Benjamini–Hochberg FDR (*** *q* < 0.001 across all pairwise comparisons).

**Figure 3:**
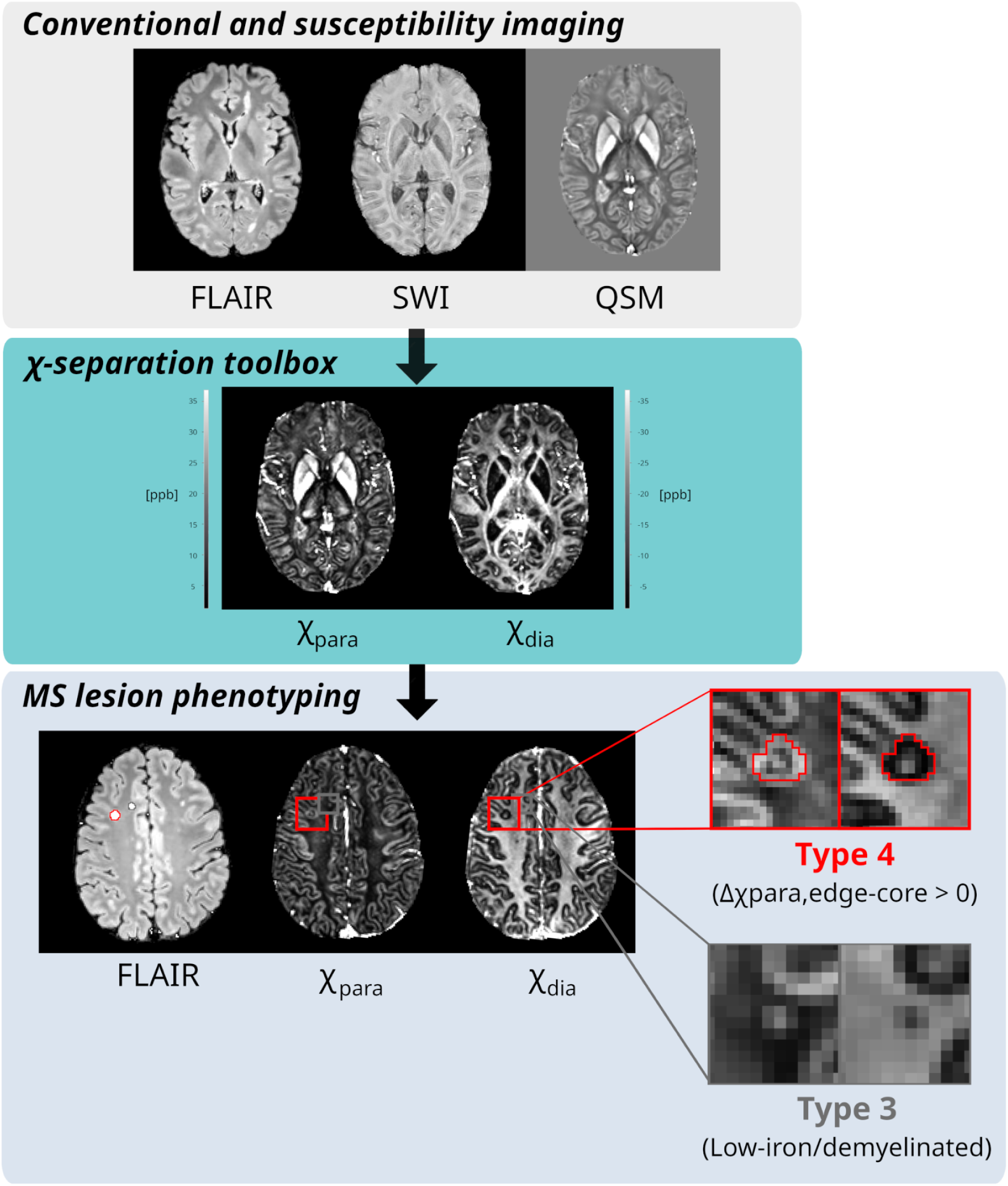
Qualitative interpretability of χ-separation phenotypes. The Type 4 phenotype (red box) captures a Paramagnetic Rim Lesion (PRL) quantitatively, exhibiting a positive paramagnetic edge-core gradient (Δχ_para,edge-core_ > 0), signifying an active peripheral border. In contrast, the Type 3 phenotype (grey box) presents as a chronically demyelinated lesion lacking paramagnetic rim with uniform susceptibility loss.

**Table 2.**
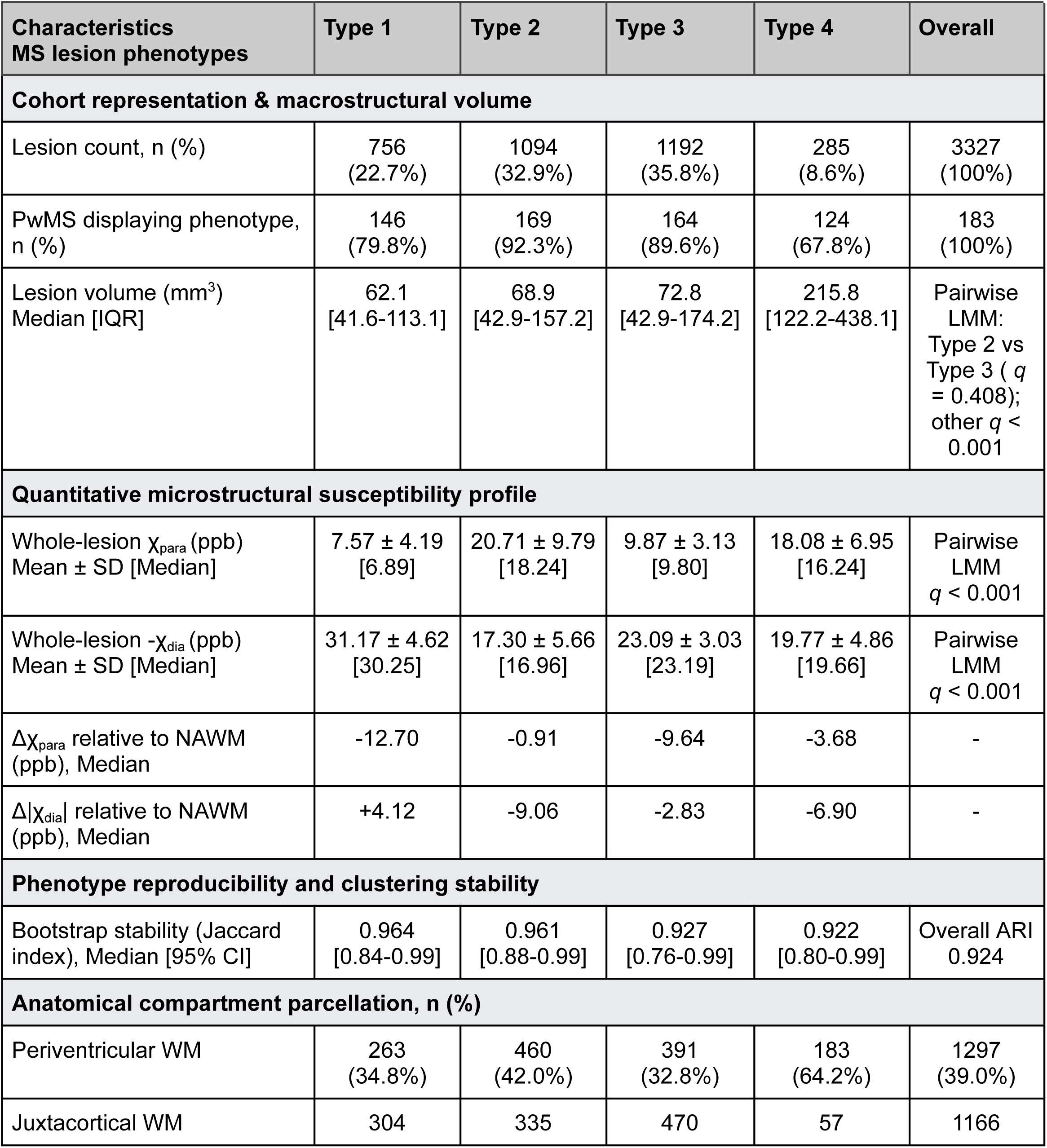
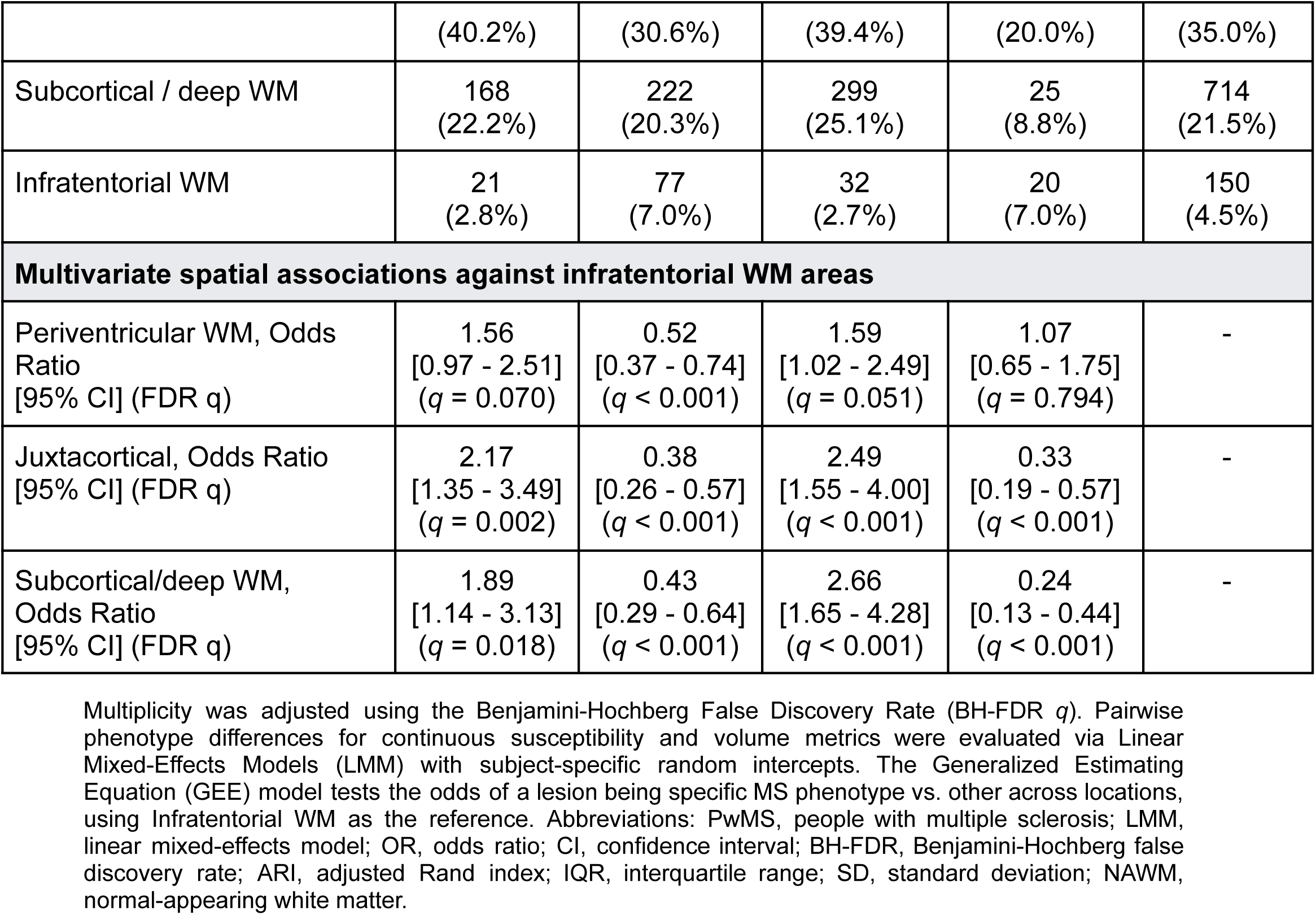
Microstructural susceptibility profiles, bootstrap reproducibility and anatomical topographies of the four MS lesions phenotypes in GRE_5_.

### 3.2 Lesion-level microstructural contrasts and anatomical topography

LMM with subject-specific random intercepts characterized the intralesional susceptibility profiles across phenotypes while accounting for within-subject lesion clustering. In addition to high partition stability (overall ARI = 0.924, 95% CI: [0.758, 0.992], with median Jaccard indices ranging from 0.922 to 0.964 across all lesion phenotypes; Table 2), the four MS lesion phenotypes exhibited distinct pairwise microstructural differences across all comparisons ( all *q* < 0.001). Furthermore, the four lesion phenotypes exhibited distinct spatial topographies across cerebral WM compartments (see Supplementary Figure 2 and Table 2). Type 1 and Type 3 lesions were significantly enriched in juxtacortical and subcortical/deep WM locations relative to infratentorial WM areas (OR ≥ 1.89 – 2.66, *q* ≤ 0.018). In contrast, Type 2 and Type 4 lesions were markedly underrepresented in both juxtacortical and subcortical/deep WM locations (OR ≤ 0.24 – 0.43, all *q* < 0.001), preferentially concentrating in periventricular regions (64.2% for Type 4 and 42.0% for Type 2) and infratentorial WM (7.0% each).

### 3.3 Independent clinical associations by MS lesion phenotypes

To establish the clinical relevance of phenotype-specific microstructural findings, multivariable linear regression models adjusted for age and sex (with separate sensitivity models additionally adjusting for subject-level total log-transformed lesion volume or total lesion count, or for DD) were evaluated with respect to EDSS and SDMT. Higher EDSS was associated with intralesional susceptibility measures within Type 4 and Type 2 lesions, whereas the low-χ_para_ Type 1 and Type 3 phenotypes demonstrated no significant independent associations with physical disability across primary or sensitivity models (see Supplementary Figure 3):

● Multivariable linear regression demonstrated that higher EDSS scores were associated with diamagnetic susceptibility alterations in the high-χ_para_ parent branch. In Type 4 lesions, higher EDSS consistently correlated with more negative signed χ_dia_ across all compartments: whole lesion (β = –0.37, *q* = 0.020), core (β = –0.37, *q* = 0.006), and edge (β = –0.34, *q* = 0.030). These associations remained significant after separate additional adjustment for total lesion volume, total lesion count, or DD. The Type 4 core association persisted in total-volume-adjusted (β = –0.35, *q* = 0.004), total-count-adjusted (β = –0.37, *q* = 0.008), and age-, sex-, and DD-adjusted models (β = –0.32, *q* = 0.033). The Type 2 edge association was significant in primary (β = –0.33, *q* = 0.044), total volume adjusted (β = –0.32, *q* = 0.030), and total count adjusted models (β = –0.35, *q* = 0.017), but did not survive FDR correction after additional adjustment for DD (*q* = 0.084). Exploratory total-burden sensitivity models also identified positive associations between EDSS and Type 2 whole-lesion χ_para_ (total volume: β = +0.19, *q* = 0.009; total count: β = +0.20, *q* = 0.017).
● In contrast, no phenotype-specific susceptibility associations with SDMT survived FDR correction in the primary age– and sex-adjusted models across all three compartments (all *q* greater than 0.05).

### 3.4 Physical disability stratification and escalation of Type 4 lesion burden

To further elucidate the clinical impact of Type 4 lesions, we stratified the cohort into three clinically defined disability stages: mild physical disability (EDSS ≤ 2.5; n = 98, 62.0%), moderate disability (EDSS 3.0–5.5; n = 36, 22.8%), and severe disability (EDSS ≥ 6.0; n = 24, 15.2%). Across disability stages, Type 4 lesion frequency and internal tissue alterations increased progressively (see Figure 4):

**Figure 4:**
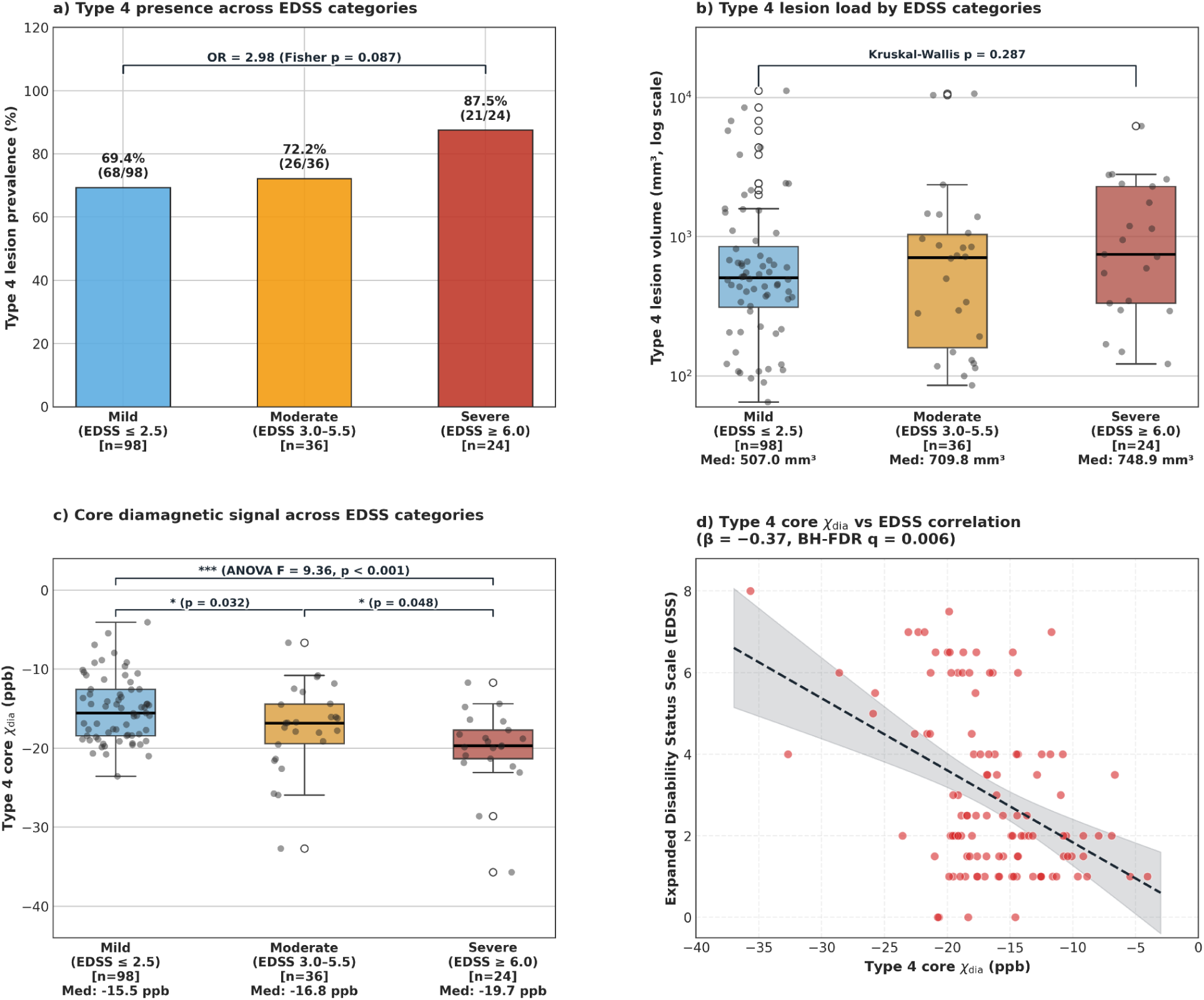
Physical disability stratification and microstructural escalation in Type 4 lesions: (a) Prevalence of Type 4 lesions across clinical disability stratification: mild (EDSS ≤ 2.5, n = 98), moderate (EDSS 3.0–5.5, n = 36), and severe (EDSS ≥ 6.0, n = 24). Type 4 lesions are identified in 87.5% (21/24) of PwMS with severe physical disability, compared to 72.2% (26/36) in moderate disability, and 69.4% (68/98) in mild disability. PwMS without Type 4 lesions (n = 43) had a lower observed frequency of severe disability although the difference was not statistically significant (only 3 of 43 [7.0%] reaching EDSS ≥ 6.0; Fisher exact test p = 0.087, Odds Ratio = 2.98). (b) Among PwMS with Type 4 lesions, median cumulative lesion volume was numerically higher across EDSS disability groups (mild: 507.0 mm³; moderate: 709.8 mm³; severe: 748.9 mm³). (c) Core diamagnetic signal escalation across disability strata. Median signed core χ_dia_ shifts progressively towards more negative values in advanced disability (mild: –15.5 ppb; moderate: –16.8 ppb; severe: –19.7 ppb; one-way ANOVA F = 9.36, p < 0.001), with group medians shown below the horizontal axis. (d) The Type 4 core association with EDSS remained significant after separate adjustment for total lesion volume (β = −0.35, *q* = 0.004), total lesion count (β = −0.37, *q* = 0.008), or DD (β = −0.32, *q* = 0.033).

#### Type 4 prevalence across disability stratification

The subject-level prevalence of Type 4 lesions was higher in groups with greater disability. Type 4 lesions were identified in 87.5% (21/24) of PwMS with severe physical disability (EDSS ≥ 6.0), compared with 72.2% (26/36) with moderate disability and 69.4% (68/98) with mild disability. Severe disability was present in 3/43 participants without Type 4 lesions (7.0%) and 21/115 participants with Type 4 lesions (18.3%). This difference did not reach statistical significance (two-sided Fisher exact p = 0.087; odds ratio (OR) = 2.98).

#### Stepwise escalation of Type 4 volume

Among PwMS with Type 4 lesions, median cumulative lesion volume was numerically higher across mild, moderate, and severe disability groups (507.0, 709.8 and 748.9 mm³, respectively) although the between-group difference was not statistically significant (Kruskal–Wallis *p* = 0.287).

#### Intralesional core diamagnetic alterations

Analysis of internal compartmental susceptibility revealed that median Type 4 core signed χ_dia_ was progressively more negative across increasing disability categories (one-way ANOVA F = 9.36, p < 0.001). Median signed core χ_dia_ shifted from –15.5 ppb (IQR: –18.4 to –12.6 ppb) in mildly disabled patients to –16.8 ppb (IQR: –19.4 to –14.4 ppb) in moderate disability, and reached –19.7 ppb (IQR: –21.3 to –17.7 ppb) in severely disabled patients. In multivariable continuous models adjusted for age and sex, Type 4 core signed χ_dia_ demonstrated a robust inverse correlation with continuous EDSS (β = –0.37, 95% CI: [-0.56, –0.18], *q* = 0.006), which remained significant after separate adjustment for total lesion volume (β = –0.35, *q* = 0.004), total lesion count (β = –0.37, *q* = 0.008), or DD (β = –0.32, *q* = 0.033).

### 3.5 MS lesion phenotype composition across disease duration and physical disability

Multivariable regressions adjusted for age and sex evaluated cross-sectional associations between DD and absolute phenotype-specific log-transformed lesion volumes and counts.

● **MS lesion phenotype over DD:** Longer DD was significantly associated with greater absolute volumes and counts of Type 1 (β = +0.33 volume, +0.34 count, both *q* < 0.001) and Type 3 lesions (β = +0.37 volume, +0.32 count, both *q* < 0.001), alongside a weaker association with Type 4 burden (β = +0.15 volume/count, *q* = 0.020). In contrast, Type 2 lesion volume and count were not significantly associated with DD (*q* = 0.441; Table 3).
● **Association with EDSS:** Subject-clustered bootstrap Spearman rank correlations identified exploratory associations, summarized using Spearman correlation coefficients and 95% bootstrap confidence intervals between macrostructural lesion phenotype composition and physical disability. Higher disability was positively correlated with Type 1 count percentage (ρ = +0.19, 95% CI: [+0.02, +0.35]) and Type 3 volume percentage (ρ = +0.18, 95% CI: [+0.02, +0.32]). Conversely, both Type 2 volume (ρ = –0.21, 95% CI: [-0.36, –0.06]) and count percentage (ρ = –0.16, 95% CI: [-0.31, –0.01]) were negatively correlated with EDSS, reflecting a lower relative Type 2 contribution among participants with higher EDSS.

**Table 3.** Multivariable clinical DD associations across the 4 MS lesion phenotypes.

| **MS phenotypes** | **Standardized $\beta$ [95% CI]** | **BH-FDR q** |
| --- | --- | --- |
| **Multivariable linear regressions against DD (N = 161)** |  |  |
| Type 1 lesion volume | +0.33 [+0.23, +0.44] | **$q < 0.001$** |
| Type 1 lesion count | +0.34 [+0.22, +0.45] | **$q < 0.001$** |
| Type 2 lesion volume | -0.05 [-0.18, +0.08] | 0.441 |
| Type 2 lesion count | -0.06 [-0.19, +0.08] | 0.441 |
| Type 3 lesion volume | +0.37 [+0.25, +0.48] | **$q < 0.001$** |
| Type 3 lesion count | +0.32 [+0.20, +0.44] | **$q < 0.001$** |
| Type 4 lesion volume | +0.15 [+0.03, +0.27] | **0.020** |
| Type 4 lesion count | +0.15 [+0.03, +0.27] | **0.020** |

Data represent standardized regression coefficients ( $\beta$ ) from ordinary least squares linear models with disease duration as dependent variable, adjusted for age and sex. Multiplicity corrected using Benjamini-Hochberg FDR.

### 3.6 Technical validation of χ-separation with echo reduction (GRE_2_)

Analytical evaluation of paired WM lesions demonstrated that QSM and susceptibility sources (χ_para_, and χ_dia_) exhibit high cross-protocol concordance between GRE_5_ and GRE_2_ (Lin concordance correlation coefficient: CCC = 0.97 for QSM, 0.97 for χ_para_ and 0.95 for χ_dia_). At the whole-lesion level, GRE_2_ showed small median paired differences relative to GRE_5_ (GRE_2_ minus GRE_5_: χ_para_ = +0.18 ppb; and displayed |χ_dia_| = −0.12 ppb). Regional analyses nevertheless identified FDR-significant susceptibility differences between GRE_5_ and GRE_2_ (*q* < 0.05; Figure 5). The regional effect sizes were calculated as GRE_5_ minus GRE_2_, with positive values indicating higher measurements under GRE_5_ and negative values indicating higher measurements under GRE_2_. These findings indicate higher cortical QSM measurements and lower χ_para_ and χ_dia_ measurements under GRE_5_ in the respective significant regions.

**Figure 5:**
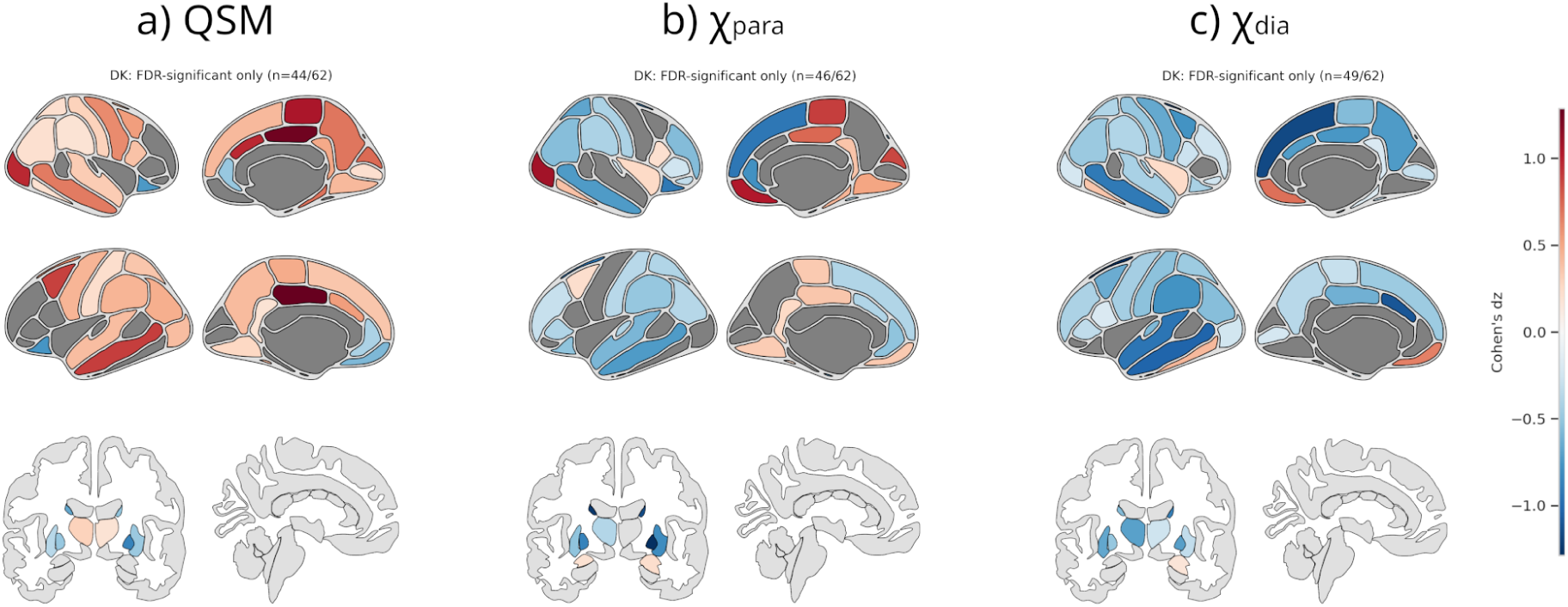
BH-FDR significant regional susceptibility differences between GRE_5_ and GRE_2_: Spatial distribution of paired effect sizes (Cohen’s dz) across the Desikan-Killiany atlas for QSM, χ_para_, and χ_dia_ (only regions surviving BH-FDR *q* < 0.05 are shown). The color scale reflects paired standardized differences between GRE_5_ and GRE_2_: Positive dz indicates higher measurements under GRE_5_; negative dz indicates higher measurements under GRE_2_. Significant differences occurred in 44/62 cortical regions for QSM, 46/62 for χ_para_, and 49/62 for displayed diamagnetic magnitude.

To evaluate the clinical translation of an abbreviated protocol, we replicated the phenotyping and clinical modeling pipeline using the GRE_2_ dataset. While GRE_2_ recovered the four MS lesion phenotypes with comparable lesion counts (p= 0.217) and preserved clinical associations within the high-χ_para_ parent branch, specifically highlighting associations in Type 2 lesions (core: β = –0.30, *q* < 0.05; edge: β = –0.42, *q* < 0.001; see Supplementary Figure 4), it exhibited distinct compartmental trade-offs. Specifically, the Type 4 core-specific association detected under GRE_5_ (β = –0.37, *q* = 0.006) was diluted under GRE_2_ (β = –0.20, *q* = 0.215).

## 4. Discussion

Our large-scale study of over 3000 quality-controlled lesions in 183 PwMS established a hybrid framework combining unsupervised clustering and hierarchical rule-based assignment to characterize the microstructural heterogeneity of MS lesions using biophysical magnetic susceptibility source separation (χ-separation). Using multi-echo GRE_5_ acquisitions, this framework distinguishes four distinct and reproducible MS lesion phenotypes (Types 1–4; ARI = 0.924) that capture divergent states of paramagnetic neuroinflammation and diamagnetic tissue integrity. Importantly, our findings contribute to understanding the clinico-radiological paradox by identifying susceptibility associations with EDSS that remained significant after separate adjustment for total lesion volume, total lesion count, or DD. Finally, by quantitatively benchmarking multi-echo protocols against international consensus guidelines, we demonstrate that while QSM remains relatively robust to sequence abbreviation in GRE_2_ (CCC ≥ 0.951), the reported compartment-specific clinical associations differed between GRE_5_ and GRE_2_.

### 4.1 Decoding the clinico-radiological paradox

Decomposed quantitative susceptibility maps provide additional insight into the classic clinico-radiological paradox^11^. Conventional T2-weighted and T2-FLAIR sequences uniformly depict edema, demyelination, gliosis and axonal loss as non-specific hyperintensities, failing to distinguish CALs from inactive chronic lesions. Furthermore, although advanced imaging biomarkers such as SELs quantify chronic radial expansion using deformation fields, they cannot characterize the underlying tissue pathology or confirm the presence of an iron-rich rim of activated microglia. Moreover, their assessment requires longitudinal MRI data. QSM provides a physical mechanism to quantify tissue composition in vivo, but represents the net volume-averaged sum of all magnetic sources within a voxel. In MS lesions, bulk susceptibility combines paramagnetic and diamagnetic contributions; therefore, similar net values can arise from different tissue compositions, limiting the distinction between iron-related and myelin-related changes.

By decomposing χ_para_ and χ_dia_ sources through χ-separation, distinct biophysical shifts that remain invisible on QSM are clearly distinguished: Type 1 lesions exhibit minimal paramagnetic accumulation coupled with the highest preserved diamagnetism, denoting relative myelin preservation^45^. On the other hand, Type 3 lesions represent a chronic demyelinating disease aspect displaying marked diamagnetic loss without an inflammatory paramagnetic footprint, aligning histopathologically with chronic inactive lesions characterized by demyelination with an absence or near absence of macrophages and activated microglia (no active inflammation)^46^. Notably, no FDR significant susceptibility associations with EDSS were detected for Types 1 and 3. These findings do not establish that these phenotypes are clinically inconsequential but explain why PwMS can accrue substantial chronic lesion volume without progressive physical disability. Most importantly, our hierarchical approach identified the Type 4 phenotype, which presented Δχ_para,edge-core_ greater than 0 ppb, as having the highest MS lesion volume compared to all other phenotypes, in line with the systematic review and meta-analysis by Tranfa M et al^9^. Furthermore, although Type 4 lesions are defined quantitatively by a positive edge-core paramagnetic gradient rather than by visual rim assessment, this spatial profile motivates comparison with rim-associated pathology, specifically mirroring the neuropathology of CALs that feature a hypercellular perilesional margin of iron-accumulating, pro-inflammatory microglia and macrophages actively driving tissue damage^5,47^. Previous studies have linked broad rim lesions to rapid disease progression and greater tissue damage within PRLs to higher disability and disease severity^48,49^. Our clinical regressions support this association by demonstrating that EDSS was associated with intralesional susceptibility alterations within Type 4 lesions after separate adjustment for total lesion burden or DD.

Importantly, Type 4 lesions were also present in 69.4% of PwMS with mild disability (EDSS ≤ 2.5), indicating that their presence alone is not specific to advanced disability. Rather, greater physical disability was associated with intralesional susceptibility characteristics, marked by a progressive escalation in diamagnetic magnitude across disability stages (shifting from –15.5 ppb in mild to –19.7 ppb in severe disability). More negative core χ_dia_ was robustly associated with higher EDSS in continuous models, and remained significant after separate adjustment for total lesion volume, lesion count or DD, suggesting that core susceptibility provides information about disability beyond total lesion burden and disease chronicity. Meanwhile, higher EDSS was associated with more negative signed χ_dia_ at the edge of Type 2 lesions, although this association did not survive FDR correction after additional adjustment for DD. In parallel, a sensitivity model adjusted for total lesion volume identified a positive association between EDSS and Type 2 whole-lesion χ_para_. One possible explanation is that Type 2 lesions represent uniformly active or resolving subacute plaques exhibiting diffuse, whole-lesion infiltration of iron-rich macrophages and microglia rather than a rim-restricted pattern^50^, consistent with pathological evidence linking altered iron distribution to oxidative injury in MS lesions^51^. Controlling for macroscopic volume isolates the intrinsic density of this diffuse inflammatory iron, whereas the attenuation of edge susceptibility with DD is consistent with the known gradual burnout and clearance of inflammatory phagocytes as active lesions mature^46^. Complementary evidence comes from the reported association between bulk susceptibility in non-ring QSM-positive lesions and EDSS^52^. Together, these findings suggest that clinically relevant paramagnetic alterations can occur in lesions without a demonstrated positive edge-core susceptibility gradient.

More negative signed χ_dia_ (reflecting greater diamagnetic magnitude) within Type 4 cores was associated with greater disability after separate adjustment for total lesion volume, total lesion count, or DD. These findings suggest that compartment-specific susceptibility provides information about disability beyond overall lesion burden. Biophysically, this direction of effect appears counterintuitive if χ_dia_ is interpreted exclusively as a linear surrogate for intact myelin content, where advanced demyelination would be expected to shift signed χ_dia_ toward zero. However, other tissue microstructural and architectural properties may complicate this direct relationship because χ_dia_ reflects other constituents such as axonal neurofilaments, glial processes and extracellular content^27^. Specifically, a recent study proposed that vasogenic edema and reactive astrogliosis, through changes in tissue water and diamagnetic protein content, could counterbalance the reduction in diamagnetic magnitude expected from demyelination^53^. Furthermore, susceptibility source separation research in Alzheimer’s disease suggests that diamagnetic component measurements may also be sensitive to protein-related pathology, supporting consideration of non-myelin contributions to diamagnetic properties^54^. In MS, postmortem spatial protein profiling has demonstrated increased GFAP-positive astrocytic process density and distinct astrocytic protein signatures within chronic active lesion cores, supporting the presence of substantial non-myelin tissue remodeling in this compartment^55^. We therefore hypothesize that severe core tissue destruction, dense fibrillary astrogliosis, or residual diamagnetic breakdown derivatives contribute to the greater diamagnetic magnitude observed in Type 4 lesion cores among PwMS with higher EDSS. Such remodeling could coexist with persistent axonal injury, providing a plausible biological explanation for the association between greater diamagnetic magnitude and disability despite an apparently stronger myelin-sensitive signal.

### 4.2 Spatial topography, disease duration and lesion phenotype composition

Anatomical parcellation demonstrated that these four susceptibility phenotypes exhibited distinct spatial topographies across cerebral WM compartments. Type 2 and 4 lesions were heavily concentrated in the periventricular space, whereas both high χ_para_ lesion phenotypes were markedly underrepresented in juxtacortical and subcortical/deep WM compartments. This relationship aligns with previously reported findings demonstrating that the expansion and destructive severity of chronic MS lesions follow a periventricular gradient, being most prominent in close proximity to cerebrospinal fluid (CSF) and declining as the distance from the ventricles increases^56^. In contrast, Type 1 and Type 3 lesions showed marked enrichment in juxtacortical and subcortical/deep WM locations. Subject random-intercept LMM confirmed significant overall location effects on lesion volume across all four lesion phenotypes, with periventricular lesions consistently displaying the largest volumes. Evaluating DD provided microstructural insight into disease chronicity. Greater DD was associated with lower χ_para_ concentrations alongside cumulative χ_dia_ loss, consistent with the neuropathological transition from active inflammation toward burned-out chronic inactive lesions^46^. The cross-sectional associations with DD do not establish whether Type 4 lesions with positive edge-core gradient convert toward lower-χ_para_ phenotypes over time. In the primary clinical models, diamagnetic susceptibility measures within Types 2 and 4 were associated with higher EDSS, whereas no FDR-significant susceptibility associations were detected for Types 1 and 3. The underlying cellular and tissue mechanisms remain uncertain. In line with this, higher EDSS correlated positively with Type 1 count percentage and Type 3 volume percentage, whereas both Type 2 volume and count percentages were negatively correlated. These cross-sectional relationships indicate a lower relative contribution of Type 2 lesions among PwMS with higher EDSS, whereas higher EDSS was associated with a greater Type 1 count percentage and Type 3 volume percentage.

### 4.3 Echo sampling and compartment-specific clinical association

The methodological comparison between GRE_5_ and GRE_2_ highlights differences in the detection of phenotype-specific associations with physical disability. The positive edge-core gradient of Type 4 lesions motivates comparison with PRLs, although correspondence with PRLs or histologically defined CALs was not established^3,5^. More negative signed χ_dia_ within Type 4 cores was associated with higher EDSS. However, the respective contributions of myelin, gliosis, and other tissue constituents remain unresolved. In contrast, Type 2 lesions exhibit higher χ_para_ with non-positive or unavailable edge-core gradients; however, their inflammatory activity and temporal evolution were not established. Notably, while abbreviated GRE_2_ maintained whole-lesion concordance, it attenuated the core-specific Type 4 clinical association to non-significance (*q* = 0.215), while clinical associations remained detectable within Type 2 lesions. The reported Type 4 core association was weaker under GRE2 and did not survive FDR correction. These findings suggest sensitivity to echo sampling, but do not establish the mechanism of the difference or demonstrate that GRE_5_ is required.

### 4.4 Methodological limitations and future directions

Current χ-separation algorithms carry several biophysical limitations. First, the optimization framework assumes isotropic susceptibility distributions. In highly oriented WM tracts (such as the corpus callosum), the magnetic susceptibility of myelin exhibits strong anisotropic behaviour, which is highly dependent on the physical angle of the fiber relative to the main B0 magnetic field^27^. Future algorithmic enhancements integrating these microstructural fiber orientation dependencies using advanced diffusion MRI will further refine the absolute precision of myelin quantification in complex WM architecture^57^. Furthermore, while recent longitudinal applications of χ-separation have proposed its utility in quantifying biologically meaningful properties^58^, caution is warranted regarding direct biological interpretations, and independent histological or external validation remains essential before confirming the susceptibility-derived iron and myelin surrogates. Second, direct MRI histopathological comparisons incorporating quantitative measures of myelin, gliosis, and tissue water are needed to establish their respective contributions to intralesional susceptibility sources^26^. Third, addressing the trade-offs of sequence abbreviation remains essential for clinical translation. While our findings demonstrate that abbreviated GRE_2_ achieves excellent macroscopic concordance at the whole-lesion level and reliably recovers MS lesion phenotypes, reduced echo sampling may increase uncertainty in R2* estimation and susceptibility reconstruction. Deep learning approaches trained on paired multi-echo datasets to regularize two-point phase inversion and denoise R2* mapping may help extract reliable intralesional χ-separation metrics from rapid two-echo acquistions, facilitating clinical translation without degrading intralesional rim-core precision. Finally, while our cross-sectional models characterize associations with disease duration, longitudinal MRI images within the same subjects are required to definitively characterize the evolutionary life cycle and transitional dynamics of lesion phenotypes.

## 5. Conclusions

Quantitative χ-separation identified four internally stable susceptibility-defined MS lesion phenotypes, revealing disability-associated tissue heterogeneity beyond total lesion burden. Higher EDSS was associated with more negative signed diamagnetic measures within Type 4 cores after adjustment for lesion burden, suggesting complex non-myelin intralesional remodeling or reactive astrogliosis that warrants histological validation. Type 4 prevalence and median volume were descriptively higher across groups with greater disability. The Type 4 core association was weaker under two-echo subsampling, suggesting that echo sampling may influence compartment-specific clinical associations. By uncoupling susceptibility sources, χ-separation characterizes disability-associated lesion microstructural heterogeneity independent of total macroscopic burden.

## Data availability

The data that support the findings of this study are available from the corresponding author upon reasonable request. Quantitative χ-separation pipeline is available at https://github.com/SNU-LIST/chi-separation and https://github.com/eloydelas/xseparation-bids (Upon Acceptance).

## Supplementary Figures

**Supplementary Figure 1.**
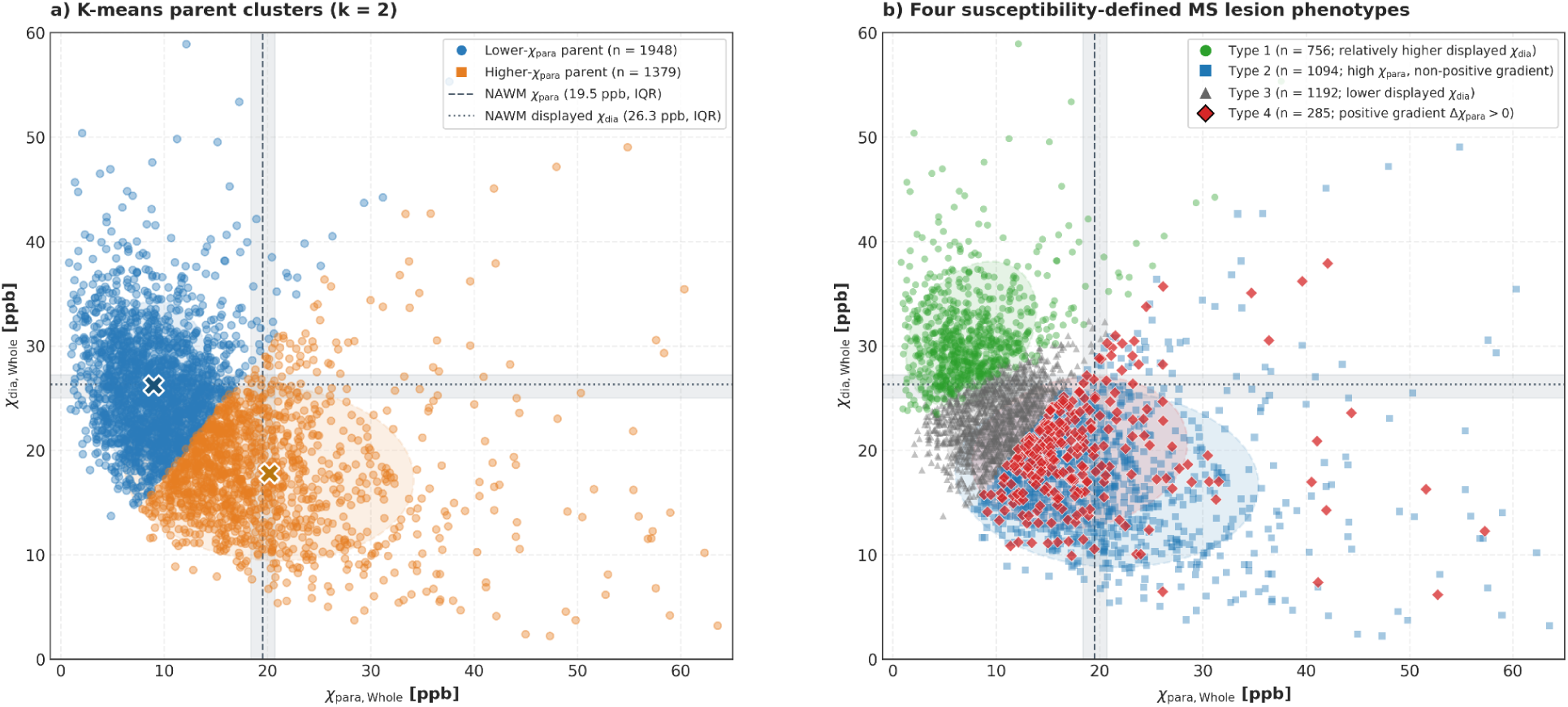
Quantitative susceptibility distribution and sequential clustering of multiple sclerosis lesion phenotypes. Scatter plot of all 3327 quality-controlled WM lesions from 183 PwMS in the whole-lesion susceptibility feature space: paramagnetic susceptibility (χ_para, whole_; x-axis) versus displayed diamagnetic susceptibility (χ_dia, whole;_ y-axis). (a) Initial unsupervised K-means clustering (k = 2) separated lesions into a lower-χ_para_ parent cluster (n= 1948, blue) and a higher-χ_para_ parent cluster (n= 1379, orange). (b) Within the lower-χ_para_ parent cluster, a second data-driven K-means step resolved lesions into Type 1 (n= 756; relatively higher displayed χ_dia_) and Type 3 (n= 1192, lower displayed χ_dia_). Within the higher-χ_para_ parent cluster, lesions exhibiting a valid positive paramagnetic edge-core gradient (Δχ_para,edge-core_>0) were identified as Type 4 (n= 285), while all remaining lesions were classified as Type 2 (n= 1094).

**Supplementary Figure 2:**
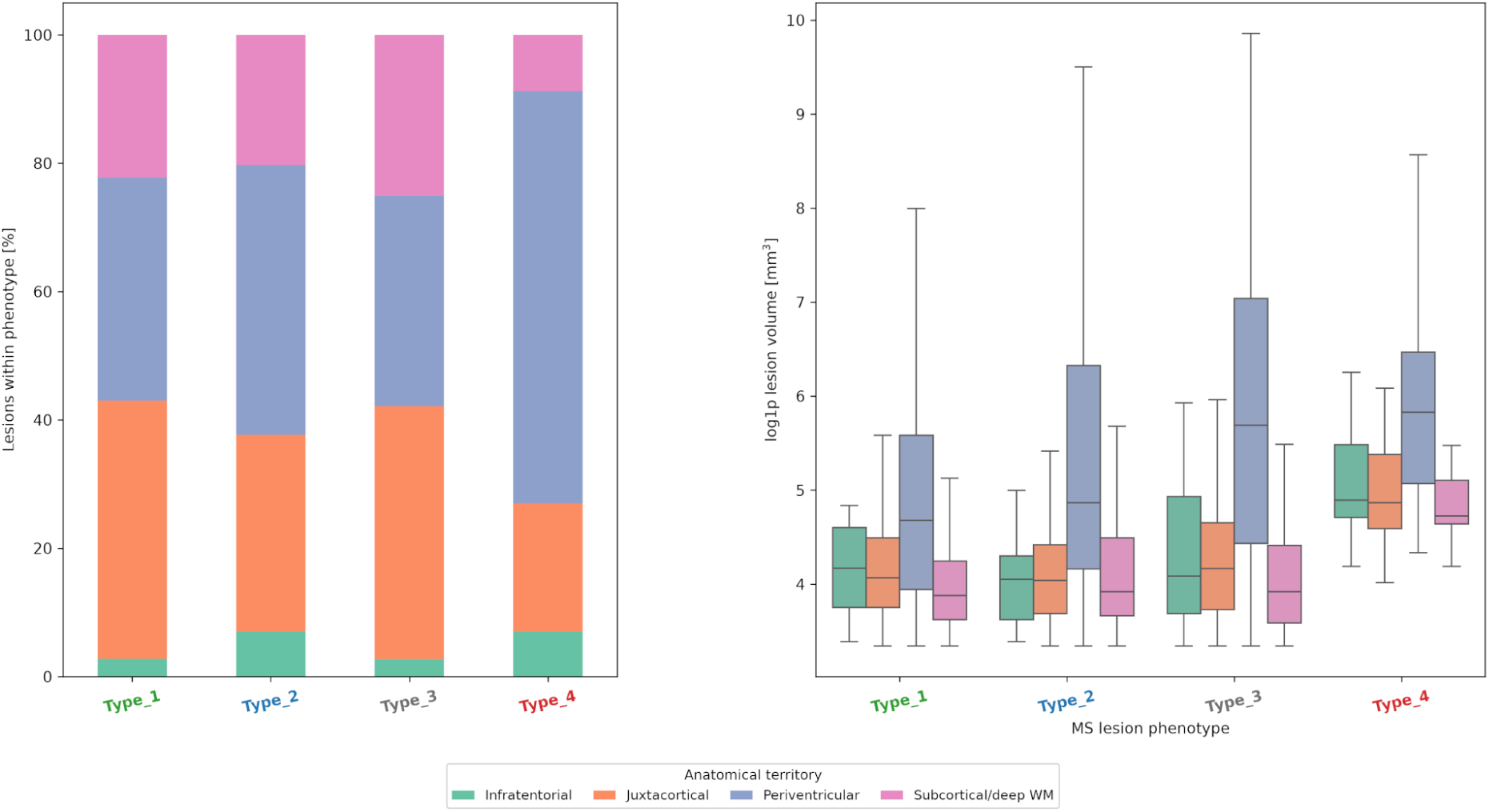
Anatomical territory composition and volume distribution across MS lesion phenotypes: (Left) Anatomical territory distribution within each phenotype. Type 4 lesions are predominantly concentrated in periventricular WM (64.2%). (Right) Log-transformed lesion volume stratified by anatomical region. Periventricular lesions consistently display the highest volumes (LMM *q* < 0.001).

**Supplementary Figure 3:**
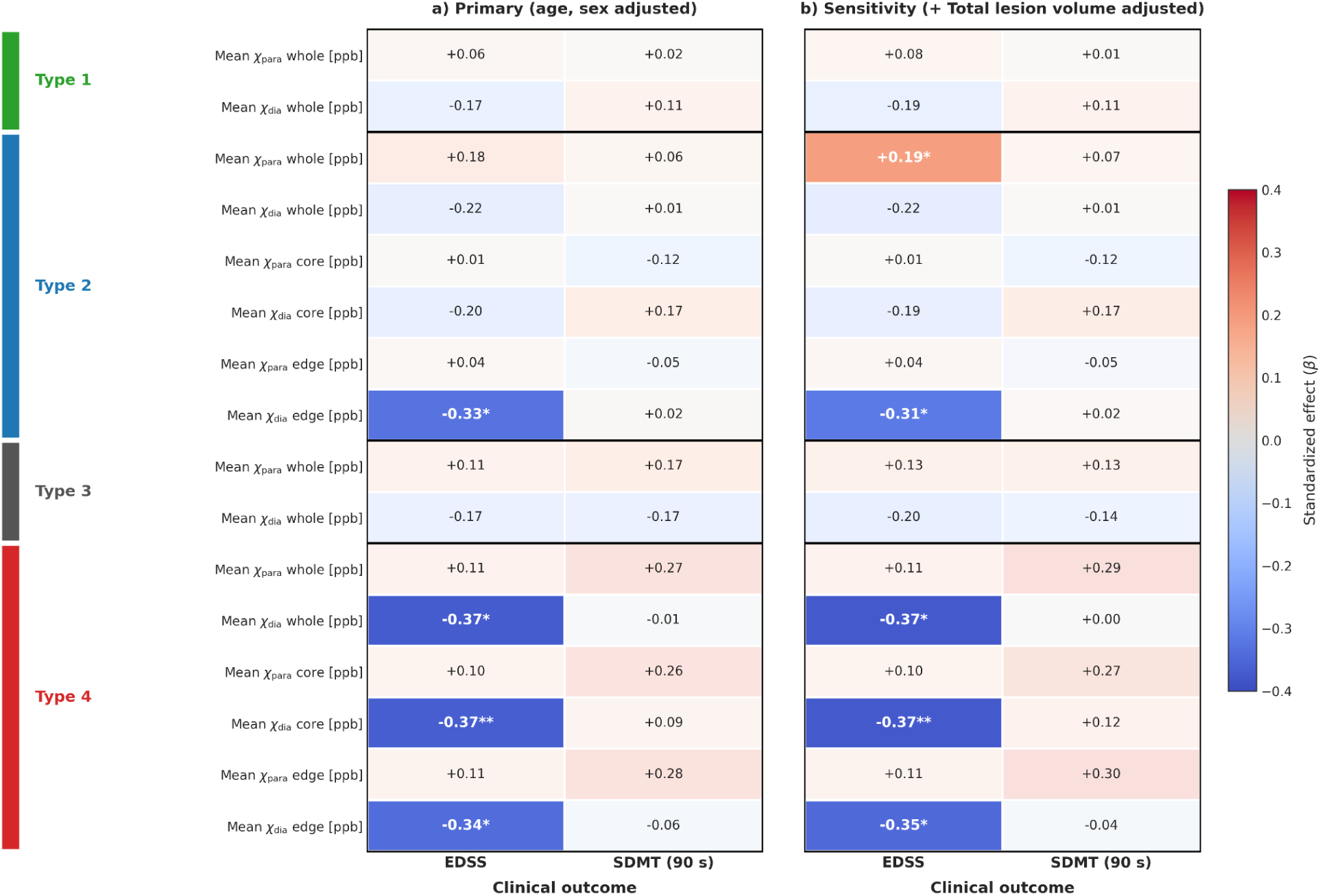
Microstructural susceptibility associations with clinical outcomes under GRE_5_: Heatmap displaying standardized regression coefficients (β) from multivariable linear models (adjusted for age and sex) evaluating physical disability (EDSS) and cognitive processing speed (SDMT) across all four MS lesion phenotypes. Intralesional concentric core-edge parcellation is methodologically and biologically grounded in the higher-χ_para_ parent branch (Types 2 and 4), where edge-core gradients define chronic rim activity (Type 4) versus uniform paramagnetic infiltration (Type 2); for low-χ_para_ phenotypes (Types 1 and 3), whole-lesion susceptibility profiles fully capture their biophysical state. Fully saturated cells indicate statistically significant associations surviving Benjamini-Hochberg False Discovery Rate correction (\**q* < 0.05, \*\**q* < 0.01). Significant associations with EDSS are exclusively localized to Type 4 (whole: β = –0.37, *q* = 0.020; core: β = –0.37, *q* = 0.006; edge: β = –0.34, *q* = 0.030) and Type 2 edge (β = –0.33, *q* = 0.044). Sensitivity models confirmed that Type 4 associations remained robust across all three compartments after further adjusting for total lesion volume: all q < 0.016; total lesion count: all *q* < 0.018, or disease duration (*q* ≤ 0.043) with the core compartment being the strongest independent predictor after separate adjustments for total lesion volume (β = −0.35, *q* = 0.004), total lesion count (β = −0.37, *q* = 0.008), or disease duration (β = −0.32, *q* = 0.033). In contrast, the Type 2 edge association lost significance after adjusting for disease duration (*q* = 0.084). Note on susceptibility sign convention: Signed χ_dia_ is mathematically defined as negative; therefore, a negative regression coefficient (β < 0) indicates that higher disability corresponds to a more negative χ_dia_ value (increased diamagnetic magnitude, with an uncertain underlying biological basis).

**Supplementary Figure 4:**
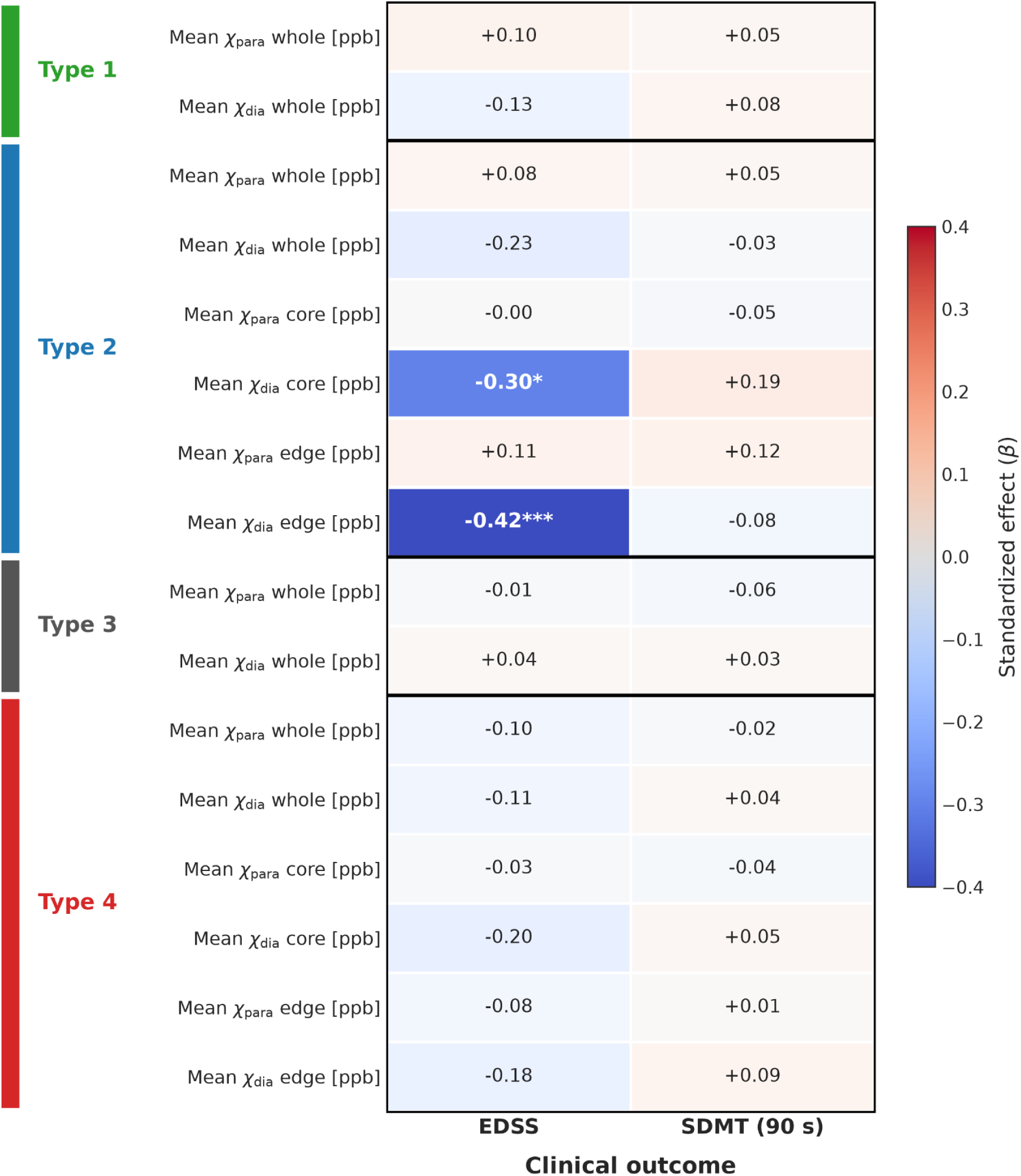
Clinical associations under the abbreviated GRE_2_ acquisition: Heatmap displaying standardized regression coefficients (β) from multivariate linear models (adjusted for age and sex) when lesion phenotypes and susceptibility metrics are derived from GRE_2_. Concentric core-edge parcellation is methodologically restricted to the high-χ_para_ parent branch (Types 2 and 4); low-χ_para_ phenotypes (Types 1 and 3) are evaluated at the whole-lesion level. Fully saturated cells indicate statistically significant associations surviving Benjamini-Hochberg False Discovery Rate correction (\**q* < 0.05, \*\*\**q* < 0.001). Under GRE_2_, the clinical relevance of high-χ_para_ subpopulation remains prominent and is concentrated in Type 2 lesions (core: β = –0.30, *q* < 0.05; edge: β = –0.42, *q* < 0.001), whereas the specific Type 4 core association is diluted (β = –0.20, *q* = 0.215 vs β = –0.37, *q* = 0.006 in GRE_5_). This illustrates that while GRE_2_ reliably captures the overall impact of paramagnetic lesion burden, multi-echo GRE5 is required to resolve subtle compartmental rim-core differences.

## Footnotes

1 Github: https://github.com/eloydelas/xseparation-bids (Upon Acceptance)

